# Determining fragility and robustness to missing data in binary outcome meta-analyses, illustrated with conflicting associations between vitamin D and cancer mortality

**DOI:** 10.1101/2025.08.15.25333793

**Authors:** David Robert Grimes

## Abstract

Meta-analysis is a vital component in clinical decision making, but previous work found binary event meta-analytic results can be fragile, affected by only a small number of patients in specific trials. Meta-analyses can also miss literature, and a method for estimating how much additional unseen data would flip results would be a useful tool. This works establishes a complementary and generalisable definition of meta-analytic fragility, based on Ellipse of Insignificance (EOI) and Region of Attainable Redaction (ROAR) methods originally developed for dichotomous outcome trials. This method does not require trial-specific alterations to estimate fragility and yields a general method to estimate robustness of a meta-analysis to data redaction or addition of hypothetical trial outcomes. This method is applied to 3 meta-analyses with conflicting findings on the association of vitamin D supplementation and cancer mortality. A full meta-analysis of all trials cited in the 3 meta-analyses yielded no association between vitamin D supplementation and cancer mortality. Using the method outlined here, it was determined that meta-analytic fragility was high in all cases, with recoding of just 5 patients in the full cohort of 133,262 patients was enough to cross the significance threshold. Small amounts of redacted or non-included data also had substantial impact on each meta-analysis, with addition of just 3 hypothetical patients to an ostensibly significant meta-analyses (N = 38,538) enough to yield a null result. This method for analytical fragility is complementary to previous investigations that suggested meta-analyses are frequently fragile. It further shows that merely increasing the sample size is not an assurance against fragility. Caution should be advised when interpreting the results of meta-analyses and conflicting results may stem from inherent fragility and should be carefully employed.

## Introduction

Meta-analyses are critical for synthesising medical evidence and providing robust estimates of treatment effects (***Murad et al., 2014***; ***Dechartres et al., 2013***; ***Nordmann et al., 2012***). However, there are concerns that meta-analyses are frequently improperly conducted, and in recent years there has been methodological concerns over the rigour of an increasing number of these publications (***Lawrence et al., 2021***; ***Siemens et al., 2021***; ***Hameed et al., 2020***), particularly when constituent Randomized Controlled Trials (RCTs) have data integrity issues which meta-analysis cannot solve (***Wilkinson et al., 2024, 2025***). There is an additional consideration that deserves mention particularly in the context of dichotomous outcome or binary outcome trials, the issue of fragility. In RCTs, many trial results are fragile, with recoding (the flipping of events to non-events or vice versa, in either or both arms) of a small number of events to non-events or vice versa in either the experimental or control arm often sufficient to convert a seemingly statistically significant result to a null, and vice versa (***Grimes, 2022***; ***Tignanelli and Napolitano, 2019***; ***Baer et al., 2021a***,b).

The issue of fragility has generated much recent discussion in RCTs, with numerous studies on various types of RCTs demonstrating that many binary results are not robust, including in oncology trials (***Del Paggio and Tannock, 2019***), cardiovascular trials (***Khan et al., 2019***), ophthalmology trials (***Shen et al., 2018***), HIV research (***Wayant et al., 2019***), and gynecologic surgery (***Pascoal et al., 2022***). FI has not however received much consideration for meta-analyses, which are often thought of as more robust than RCTs. This however may not be the case. Previously, Atal et al (***Atal et al., 2019***) suggested meta-analyses may be more fragile than might be presumed in binary outcome cases. In that work, they established an algorithm to find the minimum re-coding of patients between studies that would engineer a seemingly significant result from a null one or vice versa. Applying this to a large sample of meta-analyses, Atal et al found that many were inherently fragile. More recently, Xing et al (***Xing et al., 2025***) noted that fragility in meta-analyses was an understudied problem, and employed Atal’s method on 3758 meta-analyses to determine the mean fragility index (FI) was 5 (IQR: 2–11), with 15% of those studies having an FI of 1.

Atal’s method is highly useful, but one possible objection is that it has the downside of non-generalisability, as it finds very specific combinations of trials and patients that would have to be re-coded (events classified as non-events and vice-versa) for results to become insignificant. For example, an Atal meta-analytic fragility of 4 pertains to a specific and often unique circumstance when 4 patients could be recoded from a specific study or combinations thereof to change outputs, but this does not generalise to any 4 patients in that meta-analysis. This makes this definition of meta-analytic fragility useful but not general, and perhaps less intuitive to interpret than a typical RCT fragility metric. In this work, we establish a generalizable meta-analytic fragility metric, based upon Ellipse of Insignificance (EOI) analysis for dichotomous outcome trials (***Grimes, 2022***). This method creates a pool of events and non-events in both arms, adjusted for weighing, and answers the general question of how many patients would have to be effectively recoded in a meta-analyses for results to flip, without requiring specific study identification.

### Ellipse of Insignificance and Region of Attainable Redaction

EOI is a refined analytical fragility metric that simultaneously finds the degree of fragility in both control and experimental arms of dichotomous outcome trials. Consider any 2 × 2 chi squared test with *a* events and *b* non-events in the experimental arm and *c* events and *d* non-events in the control arm at significance level *α*. EOI theory states there is an ellipse defined by these parameters, which can be mapped on a plane with axes corresponding to the experimental control arm. It further shows that any point inside that ellipse will be non-significant at *α*, and significant when outside the ellipse. For any points outside the ellipse (significant results) then EOI analysis analytically finds and resolves the minimum displacement (Fewest Experimental/Control Knowingly Uncoded Participants (FECKUP) vector) from the ellipse, and hence true fragility of any given dichotomous outcome experiment. Conversely, if a result is inside the ellipse (non-significant), EOI will also find the minimum recoding required to reject the null. EOI is flexible,analytical, and handles samples of all size rapidly and considers changes in the control and experimental arm simultaneously, rendering it a powerful tol for robustness testing.

Intimately related to this is Region of Attainable Redaction (ROAR) analysis (***Grimes, 2024b***), a methodology for ascertaining potential influence of redacted data on reported results. There are many reasons why relevant data might be missing from an analysis, from missed studies to inappropriate cut-offs and even deliberate cherry-picking. This is highly relevant to meta-analysis, and it would be useful to have a method to quantify the potential impact of redaction on reported results and gives an additional estimate of how robust reported results may be when new data is accrued. ROAR is based on similar principles to EOI, but the bounding shape produced by the theory is no longer an ellipse, but a more complex shape. Even so, like EOI, ROAR analysis analytically and rapidly measures the displacement from the shape boundary and thus the fragility of a result to hypothetical redaction of data as well as standard re-coding in traditional fragility analysis

Both EOI and ROAR are powerful tools for 2×2 dichotomous outcome trials, and their geometric and mathematical basis has been described preciously, as well as implemented in detecting potentially spurious results recently (***Grimes and Heathers, 2025***). Accordingly in this work, we will extend these methodologies to handle meta-analysis, contrast them to Atal et al’s metric, and apply them to conflicting Vitamin D trials to illustrate their epidemiological utility. This package of meta-analytic fragility tools we name *EOIMETA* for brevity.

## Methods

### Derivation of Meta-analytic fragility and redaction analysis (EOIMETA)

For both EOI and ROAR, full mathematical details have been previously published (***Grimes, 2022, 2024b***) and an online implementation is available at https://drg85.shinyapps.io/EOIROAR/ for any dichotomous outcome trial. Extending this to the meta-analytic situation, consider *i* studies, each with an experimental group with *a*_*i*_ events and *b*_*i*_ non-events, and control group with *c*_*i*_ events and *d*_*i*_ non-events per study. If we consider for any given meta-analysis the crude unadjusted pooled relative ratio (*RR*_*p*_) and a Cochran–Mantel–Haenszel risk ratio (*RR*_*CMH*_), we can define confounding effects between studies by 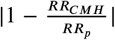. When this is small (< 10%), confounding is minimal and EOI analysis could be deployed on pooled meta-analysis results, by treating the entire meta-analysis as a single pooled sample. But as meta-analysis weigh studies by variance, simple pooling may not properly inherently account for study heterogeneity. Accordingly, we adjust using a generic inverse-variance weighed average model. For each included study, we compute relative risk and standard error as

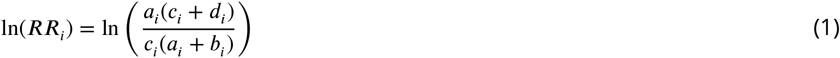

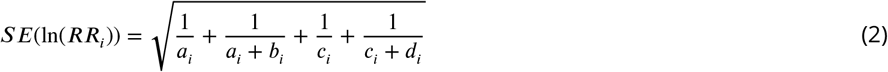

where a continuity correction of 0.5 may be applied for any zero values. The weighted average of each trial is *w*_*i*_ = *SE*(ln(*RR*_*i*_))^−1^ and total pooled risk ratio is given by

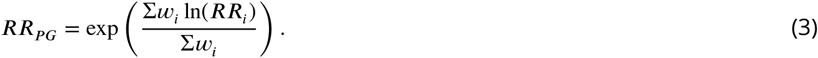

Once the pooled risk ratio is known, we can scale the event and non-event rate in the experimental group to match the adjusted relative risk while keeping the total number in the experimental arm constant. Letting *a*_*P*_ = Σ*a*_*i*_,*b*_*P*_ = Σ*b*_*i*_,*c*_*P*_ = Σ*c*_*i*_ and *d*_*P*_ = Σ*d*_*i*_, we allow 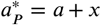 and 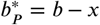 where *x* may be positive or negative to adjust the experimental group to the derived risk ratio. We can solve for this value to obtain

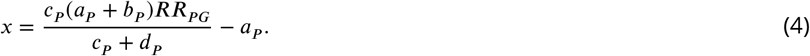

For consistency, the values of 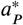 and 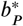 are rounded to integers. This modification in akin to pooled in a meta-analysis, and adjusts for study level heterogeneity. After this modification, a standard EOI analysis can then be applied to the vector 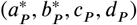. In addition, we can also employ ROAR analysis to the same vector, yielding the raw number of patients in either or both arm who could be added a given direction to change the result, and exact combination of control and experimental group redactions required to change the result from a significant finding to a null one. Caveats for implementation and interpretation are outlined in the discussion section.

### Application to vitamin D studies

Results of vitamin D supplementation and cancer mortality have been markedly inconsistent. In recent years, several meta-analyses of published trials yielded markedly conflicting results. A review and meta-analysis by Zhang et al (***Zhang et al., 2019***) and correction (***Zhang et al., 2020***) of 5 RCTs involving 38,538 patients found a significant association between vitamin D supplementation and reduced cancer mortality, reporting a 15% decrease in the risk of cancer death with vitamin D supplementation. Similarly, a meta-analysis by Gou et al (***Guo et al., 2023***) of 11 RCTs involving 111,952 patients found a significant reduction in relative risk of cancer death with supplementation of 12%. By contrast, a subsequent study of 6 RCTS involving 61,223 patients (***Zhang et al., 2022***) found no significant reduction in cancer death risk with vitamin D supplementation. All 3 meta-analyses involved combinations drawn from just 12 trials.

To investigate this apparent case of meta-analytic fragility, we extracted data from all 12 studies included in the 3 meta-analyses. A small inconsistency between all 3 meta-analysis was found in relation to one RCT (***Scragg et al., 2018***); In Zhang et al, the rate of cancer deaths in the supplementation and control group were reported as 44/2558 and 45/2550 respectively, versus being reported as 28/2558 versus 30/2558 in Gou et al and 30/2550 versus 30/2550 in Zheng et al. Disagreement between these figures appears to stem from cancers detected in both groups prior to the randomisation process. Accordingly, the adjusted ITT figure used in this work is 30/2544 versus 30/2535 respectively for Scragg et al’s work. A further nuance is that for another small RCT (***Ammann et al., 2017***), direct data was not given on the number of cases. This however can be estimated from minimal assumption through the hazard ratio. For all included studies, relative risk and 95% confidence intervals were calculated explicitly except for Lehouck et al (***Lehouck et al., 2012***) where zero recorded deaths in the supplementation sample required confidence intervals to be estimated by bootstrap methods.Initially, we ran fragility analysis on all of these meta-analysis, with the new metric outlined here as well as Atal et al’s algorithm. After this, we combined data and performed a meta-analysis on all 12 studies to ascertain potential effects of vitamin D and cancer mortality. A full fragility and robustness analysis was then performed on this, and results discussed to elucidate findings.

### Meta-analytic fragility package

We developed a custom R package (***Grimes, 2025***) (eoirroar, available at https://github.com/drg85/EOIMETA and on Zenodo) to perform the fragility and redaction analysis described in the methods, as well as Atal et al’s study specific alogorithm method for comparison.

## Results

### Meta-analytic fragility estimation of Vitamin D Cancer Mortality studies

For all conflicting meta-analyses, the unadjusted crude pooled risk ratio and CMH risk ratio were calculated prior to the analysis being conducted. For the three meta-analyses, this ranged from 0.08% − 1.1% confounding, corresponding to negligible difference. Table 1 gives the results for EOI and ROAR meta-analysis as well as the Atal et al method. Figure 2 depicts the EOI analysis results for meta-analysis by Zhang et al (including 5 RCTs, 38,538 patients) and Gou et al (11 RCTs, 111,952 patients).

**Table 1.** Application of meta-analytic fragility / redaction sensitivity to conflicting meta-analyses.

| Meta-analysis | Total patients | Risk Ratio (95% CI) | $I^2$ (95% CI) | P-value | EOI fragility | Redaction fragility | Atal fragility |
| --- | --- | --- | --- | --- | --- | --- | --- |
| Zhang et al 2019 | 38,538 | 0.85*<br>(0.74 - 0.97) | 0%<br>(0% – 79.2%) | 0.0349* | 4 | 3 | 5 |
| Guo et al 2022 | 111,952 | 0.87*<br>(0.80 - 0.95) | 0%<br>(0% – 60.2%) | 0.003* | 38 | 37 | 33 |
| Zhang et al 2022 | 61,223 | 0.96<br>(0.80 - 1.16) | 53.5%<br>(0% – 81.4%) | 0.6624 | 51 | 96 | 50 |

Figure 1 gives the EOI meta-analytic fragility vectors for both positive meta-analyses, showing the degree of recoding in both experimental and control groups required to flip results. The negative values on both axes arise because the relative risk of the experimental group (vitamin D supplementation) is ostensibly lower than the placebo group, so in the convention of EOI analysis, negative values imply additional of events to the experimental group and/or subtraction of events from the control. In the case of Zhang et al, it would take recoding of just 4 events / non-events in either experimental or control arms, or 4 recodings total in combination (< 0.01% of the total sample) to lose significance, even less than the fragility estimated by Atal et al’s algorithm. In the case of Guo et al, recoding of 38 events / non-events (< 0.038% of sample) achieves the same result, whereas there is a even more extreme specific fragility detected by Atal et al’s algorithm.

**Figure 1.**
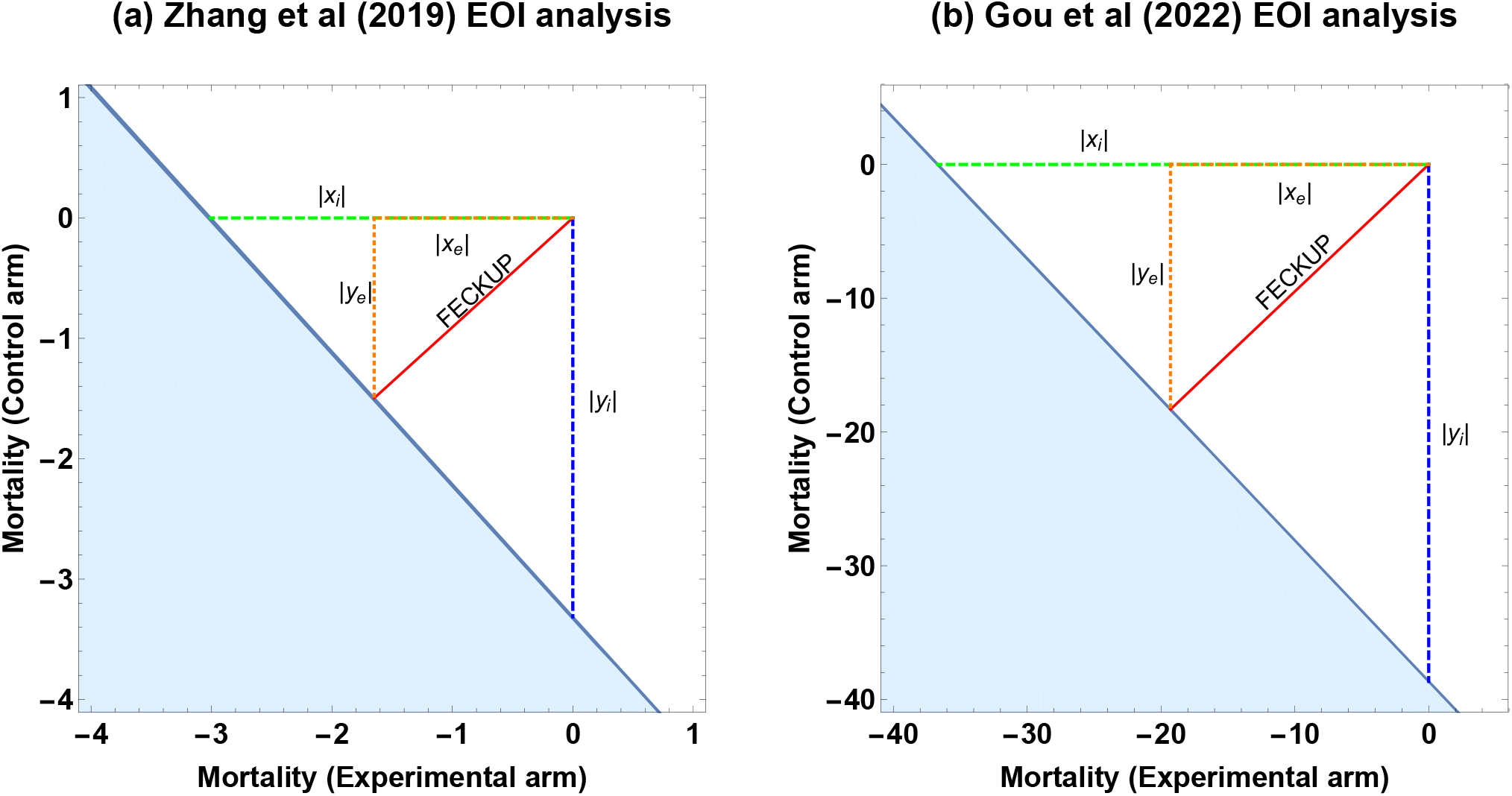
Meta-analytic EOI fragility analysis results for (a) Zhang et al and (b) Guo et al. See text for details.

The redaction analysis is also informative, because even if there were no coding or data errors in any study or their synthesis, the addition of small numbers of patients can profoundly change the result. In the case of Zhang et al, adding just 3 patients to the cohort would be enough to lose significance, while in Guo et al it is 37. Despite Zheng et al 2022 having substantially fewer patients than Guo et al 2022, it appears more robust to fragility by all metrics. This in turn leads to an important observation – the fact that a meta-analysis has more patients and studies does not inherently make it more robust or less fragile, as discussed in the next section.

### Meta-analysis of all studies

Figure 2 depicts the meta-analysis forest plot of all 12 RCTs of 133,262 patients using a random effects model, while table 2 gives summary meta-analytic fragility statistics. Table 2 gives the statistics for when all 12 studies are combined, finding that both EOI fragility and Atal et al’s fragility actually decrease despite the increase in the number of subjects. Even with a large sample, the recoding of just 5 non-events to events, in this case in the placebo arm (additional deaths in the non-vitamin D group), would alter the results to cross the threshold of significance (p = 0.048) and likely change inferences. Table 3 gives both fragility estimates for each included study on an individual basis for comparison. Note that traditional the fragility index is not defined for non-significant results, which EOI fragility is sufficiently flexible to find the counts needed to flip significance, including recoding required to move from non-significant to significant.

**Figure 2.**
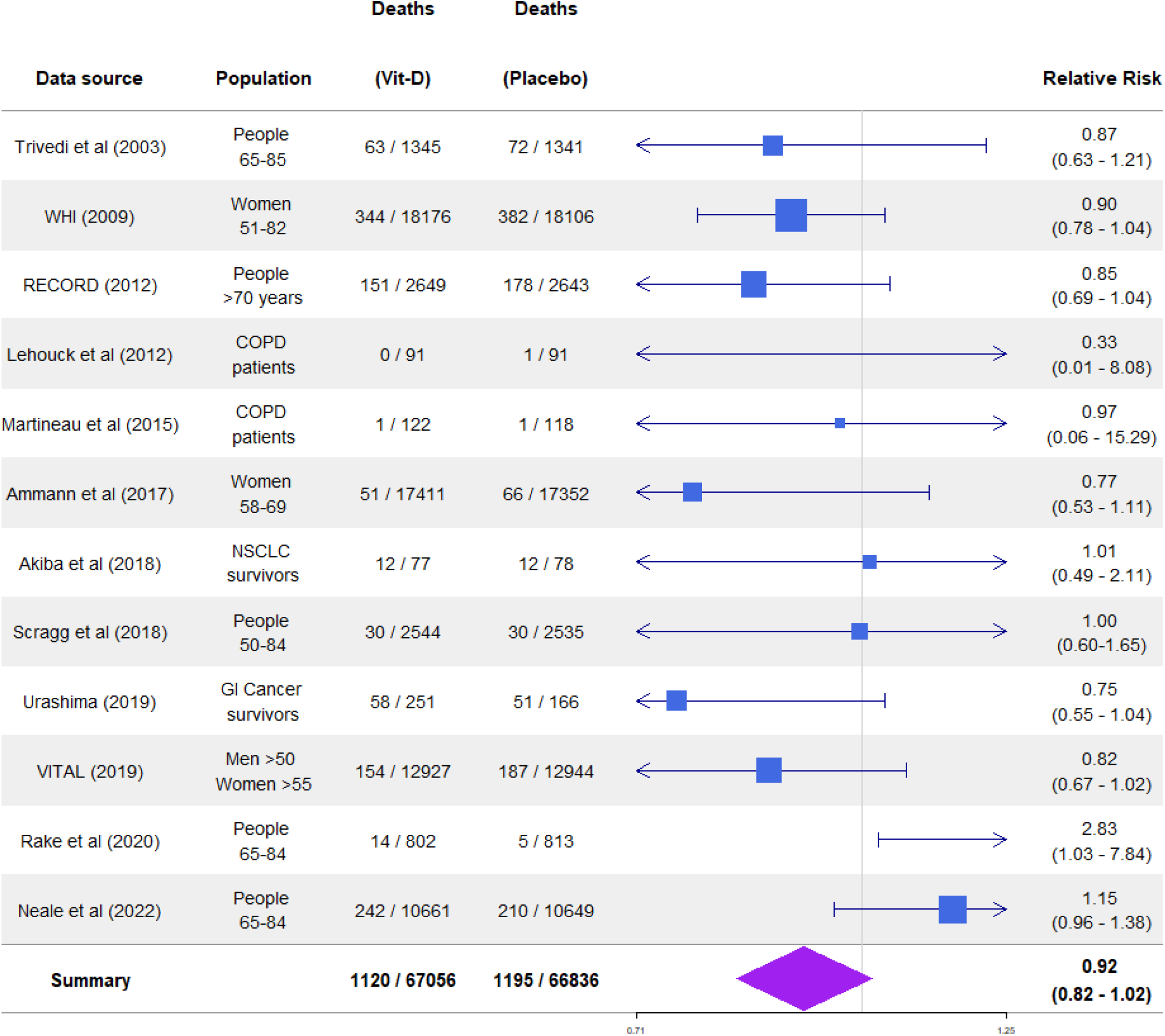
Meta-analysis of all 12 RCTS including study details and relative risk of vitamin D supplementation on cancer mortality, shown with 95% confidence intervals.

**Table 2.** Meta-analytic fragility of all 12 studies.

| Total patients | Risk Ratio (95% CI) | $I^2$ (95% CI) | P-value | EOI fragility | Redaction fragility | Atal fragility |
| --- | --- | --- | --- | --- | --- | --- |
| 133,262 | 0.92<br>(0.83 - 1.02) | 24.9%<br>(0% – 61.6%) | 0.061 | 5<br>(0.00375%) | 192<br>(0.14%) | 9<br>(0.00675%) |

**Table 3.** Fragilty comparison across all 12 studies (study-level metrics)

| Study | Deaths (Vitamin D) | Deaths (Placebo) | Fragility Index<sup>†</sup> | EOI fragility |
| --- | --- | --- | --- | --- |
| Trivedi et al (2003) | 63 / 1345 | 72 / 1341 | NA | 13 |
| WHI (2009) | 344 / 18176 | 382 / 18106 | NA | 13 |
| RECORD (2012) | 151 / 2649 | 178 / 2643 | NA | 8 |
| Lehouck et al (2012) | 0 / 91 | 1 / 91 | NA | NA<sup>λ</sup> |
| Martineau et al (2015) | 1 / 122 | 1 / 118 | NA | NA<sup>λ</sup> |
| Ammann et al (2017) | 51 / 17411 | 66 / 17352 | NA | 6 |
| Akida et al (2018) | 12 / 77 | 12 / 78 | NA | 9 |
| Scragg et al (2019) | 30 / 2544 | 30 / 2535 | NA | 15 |
| Urashima (2019) | 58 / 251 | 51 / 166 | NA | 2 |
| VITAL (2019) | 154 / 12927 | 187 / 12944 | NA | 12 |
| Rake et al (2020) | 14 / 802 | 5 / 813 | 1 | 1 |
| Neale et al (2022) | 242 / 10661 | 210 / 10649 | NA | 10 |

<sup>†</sup> Traditional FI not applicable when results are non-significant.
<sup>λ</sup> EOI Fragility ambiguous (multiple solutions) with zero or unity event counts

## Discussion

The meta-analytic fragility tool presented here, EOIMETA, is generalisable and flexible, complementing Atal et al’s study and patient specific method. The nuance between both approaches is that in Atal et al’s algorithm identifies a specific combination of patients from specific studies that may be moved to alter results, whereas the current work establishes a method for pooling meta-analysis on ascertaining their group fragility and the potential impact of missing data. In this respect, they answer complementary and subtlety different questions specific to meta-analyses.

In RCT literature, a frequent objection to the use of fragility analysis methods is that the mere existence of a small FI might be an artefact of trial design. For clinical trials especially, a well-designed trial should be structured as to minimize patient exposure to unknown harms, striving to ensure that just enough participate to allow the detection of clinically relevant effects. The argument that RCTs at least might be fragile ‘by design’ is however convincingly countered by other authors, who find no evidence of p-value distributions clustering around significance thresholds after a sample size calculation and find additional that fragility in well-designed studies is not always low. More recent robust methods for fragility analysis like EOI and for redaction like ROAR also have application far beyond ostensibly fragile-by-design RCTs, but for cohort studies, preclinical work, and even ecological studies. The fragile-by-design argument is also inherently weaker in the context of meta-analysis, where the marshalling of many studies to increase effect precision measurements should strengthen the evidence but results here and previously suggest that even meta-analysis of large groups of patients can be profoundly fragile.

There are a few subtle issues and limitations to consider additionally; Atal et al’s meta-analytic fragility metric in principle identifies the precise combination of inter-study and patient recoding required to flip a result, a “worse-case” scenario, identifying the most fragile point in a meta-analyses. This usually results in the Atal et al fragility metric being typically smaller than the complement EOI fragility metric outlined here, but this is not always the case. Because Atal et al’s method is a greedy algorithm, it is time-consuming to run on large collections of studies and can sometimes miss optimal and smaller solutions. Secondly, the adjustment of the events and non-events in the experimental group due to weighed inverse variance methods in the EOI fragility method subtly alters group composition, resulting in a smaller EOI fragility than a crude pooled estimate. What is evident from the examples shown in this work is that even in large cohorts with relatively low heterogeneity, the impact of recoding only miniscule numbers of patients can drastically alter interpretation; in the 12-study meta-analysis, recoding a mere 5 patients in 133,262 was sufficient to alter the result from a null to a positive finding.

While the method outlined here is analytic and rapid, it has some limitations that must be considered. Like Atal’s method, it applies only to meta-analyses of trials with dichotomous outcomes. It should be noted there are also fragility indices designed for continuous data (***Caldwell et al., 2021***) but these are beyond the scope of the current work. While EOIMETA has a generic weighed inverse variance (WIV) correction by default, it may not be reliable in the presence of high between-study heterogeneity. Accordingly, careful application is required. At its most essential, this implementation reflects the meta-analytic relative risk by generating a synthetic pooled 2×2 table that modifies pooled events (and non-events) in the experimental arm so that trial results may be pooled and meta-analytic fragility estimated. One interesting feature of the current method is that neglecting WIV correction and simply pooling trials for basic EOI or ROAR analysis may be justifable in some instances, especially when *RR*_*P*_ ≈ *RR*_*CMH*_ . The main effect of this simpliciation is that it produces much more fragile estimates than those presented here; for example, a simple pooling EOI analysis on Guo et al produces an EOI of 17 (as opposed to the 38 reported here) and a ROAR of just 16 (versus the 96 reported here). Thus the results reported here are thus conservative, though the precise interpretation of these variants in implementation is an open question.

The meta-analytic fragility metric is useful for modeling perturbations in results, such as the addition of patients (who were previously redacted) or the recoding of events and non-events. The advantage of this approach lies in its tractability, offering a simpler alternative to working with multiple studies individually and a deterministic and analytic resultant fragility metric. Because the resultant metric pertains to the effective pool of patients in the meta-analysis, it does not identify individual studies that may unduly influence results. For this purpose, leave-one-out analysis (***Willis and Riley, 2017***) or extensions (***Meng et al., 2024***) are more suitable tools, complementing the method described in this work.

The use of vitamin D meta-analyses in this work was chosen as illustrative rather than specific, but it is worth noting that there are methodological concerns with much Vitamin D research (***Grimes et al., 2024***). The three studies cited in this work report relatively low heterogeneity in their meta-analysis in both effect sizes and *I*^2^ values, but it is worth noting that the included studies addressed very different populations, including patients with Chronic Obstructive Pulmonary Disease, Non-small cell lung cancer survivors, women only cohorts, older adults, and gastrological cancer survivors. These groups have presumably different risk factors for cancer deaths, and why the authors of these studies combined the cohorts with fundamentally different clinical contexts is unclear. Why the heterogeneity appeared so relatively low in different groups is also a curious feature. This goes beyond the scope of the current work, but serves as an example of the reality that meta-analysis is only as strong as its underlying data and methodological rigor in comparing like-with-like, and the conclusions drawn from them must always be seen in context.

This work also raises the possibility of whether there’s an analog of “effect size” we might extend to such fragility analysis, a measure of how robust a result may be, and how much it can withstand recoding. This is a complex question in some instances, because in RCTs studies may be fragile be design, limited by trial enrollment limitations. Although this should be less of an issue in large observational trials, the Vitamin D research illustrated here suggests that only tiny fractions of the trial population (typically << 0.01%, as in table 2) would have been recoded to flip results. Whether this is typical of all types of experimental data is unclear; we can for example look at the Cohen’s *h* in this instance where 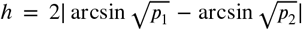 and *p*_1_ and *p*_2_ are the mortality proportion in the experimental and control group respectively. In this instance for all 12 trials, *h* ≈ 0.009. This (and the respective *h* values for all constituent meta-analyses considered) is far below the threshold of *h* = 0.2 that Cohen suggested might reflect even a ‘small’ effect, and appears virtually negligible, a weakness which might explain the fragility of the results used in this in this illustration, and it may be worthwhile to apply such a methodology to other areas in future to determine widely applicable thresholds for robustness.

While meta-analysis is a powerful method for refining effect size estimates, it cannot overcome poorly conducted research or bad data (***Jané et al., 2025***), nor it is intrinsically robust. These limitations should be kept in mind when considering meta-analytic results, and scientists should consider them doubly when opting to perform a meta-analysis. Mass produced and unreliable meta-analyses are a recognised and growing problem (***Ioannidis, 2016***), and we need be mindful not to add to increasingly issue of research waste and unreliable research (***Glasziou and Chalmers, 2018***; ***Grimes, 2024a***).

## Data Availability

All relevant code for this undertaking including the study data from the included trials is available at the linked Github repository and the Zenodo DOI

https://zenodo.org/records/15878923

## Data and code availability

All relevant code for this undertaking including the study data from the included trials is available at the linked Github repository and the Zenodo DOI, as well as additional EOI and ROAR plots stemming from this work. This article was previously uploaded as a preprint to Medrxiv (doi: 10.1101/2025.08.15.25333793).

